# Serological Markers Predict *Plasmodium vivax* Relapses in Returning Indonesian Soldier Cohorts

**DOI:** 10.64898/2026.06.08.26355218

**Authors:** Rintis Noviyanti, Retno A. S. Utami, Lauren Smith, Leily Trianty, Lenny L. Ekawati, Edwin Sutanto, Ristya Amalia, Aisah Resti Amelia, Maulina Aini Hafidzah, Nadia Fadila, Agatha M. Puspitasari, Fahira Ainun Nisa, Hidar Hidar, Pinkan P. Kariodimedjo, Aliva N. Farinisia, Gladis Hutahaean, Michael Christian, Tanti A. Kesuma, Decy Subekti, Saraswati Soebianto, Fitria Wulandari, Nunung Nuraini, Waras Budiman, Yogi Ertanto, Muhammad D. Widiartha, Furkan Furkan, Narimane Nekkab, Ramin Mazhari, Michael T. White, Leanne J. Robinson, Rhea J. Longley, J. Kevin Baird, Ivo J. Mueller

**Affiliations:** Eijkman Research Center for Molecular Biology, BRIN, Indonesia; Exeins Health Initiative, Jakarta, Indonesia; Walter & Eliza Hall Institute of Medical Research, Parkville, Australia; Oxford University Clinical Research Unit, Faculty of Medicine, Universitas Indonesia, Jakarta, Indonesia; Army Medical Center, Army of the Republic of Indonesia, Jakarta, Indonesia; Swiss Tropical and Public Health Institute, Allschwil, Switzerland; Infectious Disease Epidemiology and Analytics Unit, Department of Global Health, Institut Pasteur, Université Paris-Cité, INSERM U1347, Paris; Burnet Institute, Melbourne, Australia; Melbourne School of Population & Global Health, University of Melbourne, Australia; Department of Medical Biology, the University of Melbourne; Faculty of Tropical Medicine, Mahidol University, Bangkok, Thailand; Oxford University Clinical Research Unit, Ho Chi Minh City, Vietnam; Centre for Tropical Medicine and Global Health, Nuffield Department of Medicine, University of Oxford, United Kingdom; School of Global Health, Faculty of Medicine, Shanghai Jiao Tong University, Shanghai, China

## Abstract

**Backgroun:** Persistent transmission from relapsing *Plasmodium vivax* infections threatens malaria elimination programs in the Asia-Pacific and Americas. Tools to identify people at risk of relapse are urgently required. We aimed to validate a panel of eight *P. vivax* serological biomarkers for predicting future relapses.

**Methods:** In this observational study, soldiers returning from malaria-endemic Papua to non-endemic East Java, Indonesia, were screened at enrolment using antibody measurement (Luminex) and trained random forest classification algorithms, then followed for 6 months. Active case detection was performed fortnightly by microscopy. Algorithms classified soldiers as recently infected (last nine months) and thus at risk of relapse, based on anti-vivax antibody measurements at enrolment.

**Findings:** Between December 2018 and July 2022, 592 soldiers were enrolled, with 553 completing follow-up; 119 experienced a *P. vivax* relapse. Of these, 102 were correctly classified as at risk of relapse at enrolment, corresponding to 86% sensitivity and 86% specificity, with an AUC of 0.92.

**Interpretation:** *P. vivax* serological biomarkers can identify people at risk of relapse with high sensitivity and specificity and could be used as a novel public health intervention, *P. vivax* serological testing and treatment (PvSeroTAT), to reduce relapse-driven transmission.

**Funding:** Bill and Melinda Gates Foundation (OPP1180981) funded the study trial. LJR, RJL and IM received National Health and Medical Research Council Fellowships (#2017630, #1173210, #2016726). RJL receives salary support from the Victorian Government as a veski FAIR Fellow and from the Sylvia and Charles Viertel Charitable Foundation as a Viertel Senior Medical Research Fellow. LJR, RJL and IM receive funding from NHMRC Synergy grant (NHMRC #2018654) and are part of the Australian Centre of Research Excellence in Malaria Elimination (NHMRC #2024622).

**Conflict of interest:** IM, MTW and RJL are named inventors on a patent describing *P. vivax* serological exposure markers (PCT/US17/67926). All other authors declare no competing interests.

**Research in context:** *Evidence before this study:* Relapses from long-lasting liver stages (i.e. hypnozoites) account for 80-90% of all *P. vivax* blood-stage infections^1,2^ and it has been well understood that regional *P. vivax* elimination will be challenging without directly attacking this hidden parasite reservoir. Doing this efficiently will require novel diagnostic tests that can detect individuals at risk of relapse (as described in a recently published WHO preferred product characteristics (PPC)^3^. None of the current malaria diagnostic tests can detect hypnozoite carriers and/or identify people at risk of relapse. A previous study showed that antibodies to a validated panel of 8 *P. vivax* antigen markers can detect *P. vivax* infection within the previous 9 months with 80% sensitivity and 80% specificity, and identify people with significantly higher risk of experiencing future *P. vivax* infections.^4^ These findings suggested that these serological exposure markers (SEMs) may be a proxy for the presence of hypnozoites in the liver and thus act as biomarkers for future relapse risk.

*Added value of this study:* By following two cohorts of soldiers between 2018 to 2021, the present study has been able to confirm that antibodies to 8 *P. vivax* SEMs are able to identify individuals who experienced a *P. vivax* relapse during 6 months following their return from a malaria endemic region in Papua, Indonesia to their non-endemic base in Java with high performance (86% sensitivity, 86% specificity, AUC: 0.92). This proves that *P. vivax* SEMs are indeed markers of future relapse risk and thus valid surrogate markers for hypnozoite carriage.

*Implications of all the available evidence:* The validated *P. vivax* sero-diagnostic assay is the first ever diagnostic to fulfil the requirements outlined in the recently published WHO PPC for tests to identify individuals at risk of *P. vivax* relapse. This makes the test suitable for use in both public health interventions, such as serological testing & treatment (PvSeroTAT), where serology is used to identify people at risk of relapse to guide radical cure, and for sero-surveillance for risk stratification and monitoring and evaluation of ongoing elimination programs. Trials of both uses are currently ongoing in different vivax endemic areas of the Asia-Pacific, South America and Africa.

## Introduction

Over the last 20 years, the Asia-Pacific and the Americas have seen dramatic reductions in malaria burden^5,6^, with 26 countries achieving malaria elimination between 2000-2024^7^, and almost all other countries setting formal targets to eliminate malaria within the next 10 years. Where malaria transmission remains, *Plasmodium falciparum* transmission has often been greatly reduced, and several countries are closing in on *P. falciparum* elimination, leaving *P. vivax* as the most common infection and key challenge to achieving complete malaria elimination outside Africa.^7^ Indonesia provides a clear example of this trend. With more than one million clinical cases annually, Indonesia contributes to approximately 25% of all malaria cases in the WHO South-East Asia Region, nearly half of which are due to *P. vivax*.^7^ Although malaria remains highly endemic in eastern Indonesia, particularly Papua, substantial progress has been achieved nationally, with 389 out of 514 districts reporting zero locally acquired cases in 2023^8^, including the majority of districts in western and central Indonesia. Outside Papua, *P. vivax* now represents the dominant species and a major obstacle to elimination.

Unique features of the *P. vivax* lifecycle make it challenging to control and eliminate.^9,10^ This includes the high prevalence of asymptomatic infections, the high proportion of parasite biomass in the spleen and bone marrow, and low parasitaemia, all of which make *P. vivax* infections difficult to diagnose. However, *P. vivax* is particularly challenging due to the development of dormant hypnozoite stage parasites in the liver, which are undetectable with current diagnostic tools. After a primary blood-stage infection, hypnozoites can cause recurrent blood-stage infection (i.e. relapses) weeks to months after the initial infection. It is estimated that around 80% of all blood-stage *P. vivax* infections are due to relapses^1,2^. As such, identifying and targeting this hidden hypnozoite reservoir is essential for achieving *P. vivax* elimination. The lack of diagnostic tools capable of identifying this hidden reservoir continues to impede local elimination efforts. This has led the World Health Organisation to publish the first preferred product characteristics for diagnostics that can detect the risk of relapse.^3^

One potential approach in detecting the risk of relapse is antibody-based serology. Although there are no known biomarkers for the presence of hypnozoites in the liver, blood-stage *P. vivax* infections will induce strong IgG antibody responses towards several *P. vivax* proteins. Recently, a panel of serological markers and an associated machine learning algorithm has been developed that can classify individuals as having had a recent *P. vivax* infection in the previous 9 months with high performance.^4^ All vivax parasites, except some belonging to the hibernans strains now restricted to the Korean peninsula^7^, produce a primary blood-stage infection within 7-15 days after exposure to an infected mosquito bite, with most infected individuals relapsing within less than six to nine months after an initial blood-stage infection.^11^ Therefore, individuals who have had a blood-stage infection in the prior nine months and have not received liver-stage parasite treatment are likely to be hypnozoite carriers. This sero-diagnostic method^4^ has been able to predict risk of future *P. vivax* infections, but explicitly testing its ability to predict risk of hypnozoite relapse has been difficult due to the inability to distinguish between these and new mosquito bite-induced infections in regions with ongoing transmission.

Our study set out to assess the ability of a validated panel of *P. vivax* serological exposure markers (SEMs) to predict risk of (future) relapses. Malaria endemicity varies across regions in Indonesia: Papua has one of the highest malaria transmission rates, including *P. vivax*, with an Annual Parasite Incidence (API) of 400^12^; while East Java is certified as malaria-free, together with DKI Jakarta, Banten, Bali, and West Java.^13^ Thus, Indonesia provides an ideal setting to validate the previously developed serological markers by following naïve soldiers for 6 months after their return to malaria-free East Java, following a 9-month deployment in the malaria-endemic region of Papua. The soldiers, who were potentially exposed to *P. vivax* during deployment, remained at risk of relapse after their return to Java. Given the absence of local transmission at the home base, any malaria episodes occurring during follow-up were most likely attributable to reactivation of latent hypnozoites rather than new infections. This unique setting thus allows for direct determination of the performance of *P. vivax* SEMs for detecting individuals at risk of relapse.

## Methods

### Study design

This prospective study evaluated a panel of SEMs combined with a random forest classifier to predict relapse risk in two cohorts of Indonesian soldiers. In total, 592 male soldiers from two battalions were recruited after returning from nine-month deployment in malaria-endemic Papua to malaria-free East Java (Figure 1). Cohort one (n=294) deployed from Surabaya to Boven Digoel was followed up from December 2018 to June 2019, and Cohort two (n=298) from Malang to South Keerom was followed up from January to July 2022. Malaria episode data during deployment were unavailable, although soldiers may have been naturally exposed to *P. vivax*.

**Figure 1:**
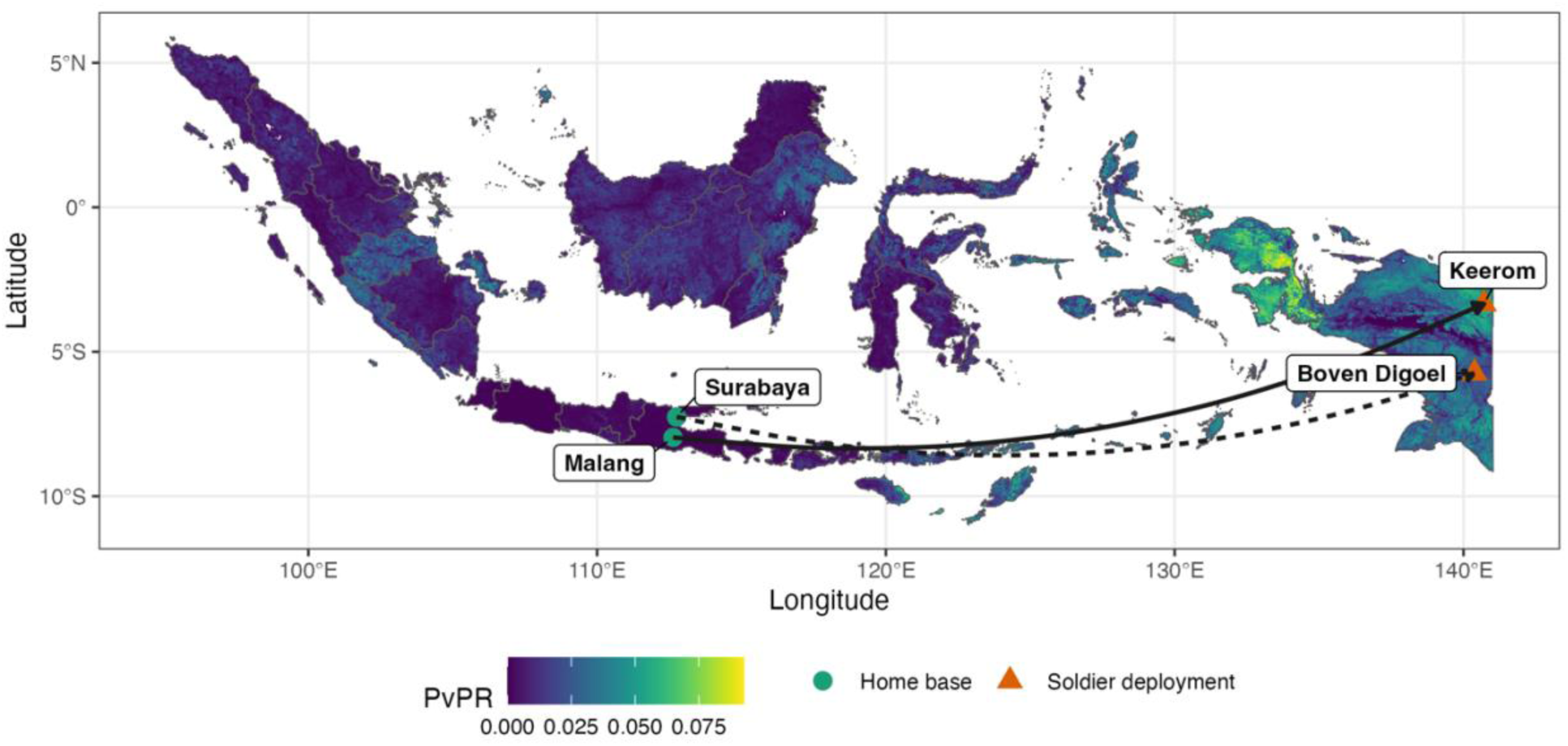
*Plasmodium vivax* prevalence (PvPR = predicted parasite rate) in Indonesia (2020) with location of soldiers during deployment (orange triangles; Keerom, Boven Digoel) and at their home base for study enrolment (green circles; Surabaya, Malang). Prevalence data were obtained from the Malaria Atlas Project^14^, using the malariaAtlas R package.

Eligible participants were aged >=18 to <65 years, had spent more than one month in Papua in the previous 12 months, and weighed 40 to 100 kilograms. Participants who required hospitalisation, had haemoglobin concentration <7 g/dL, and who planned to be off-base for 30 consecutive days were excluded.

At enrolment, blood samples were collected from all soldiers. Those *P. vivax* positive by light microscopy were treated according to national standards and not followed further. Soldiers negative at enrolment underwent active case detection (ACD) every two weeks for six months, and passive case detection (PCD) during episodes of febrile symptoms. All blood samples were screened by light microscopy to identify *P. vivax* parasitaemia and therefore relapse. Confirmatory RT-PCR was performed on all microscopy-positive samples as well as on all microscopy-negative follow-up samples from all seropositive and 20% of randomly selected seronegative individuals. Antibody measurements were taken from enrolment samples.

Written informed consent was obtained. Refusal to participating did not affect military duties, access to healthcare, or remained in the battalion. Participants were excluded from analysis if they were PCR-positive for *P. falciparum* (n=1) at enrolment or missed two or more consecutive ACD visits without experiencing relapse (n=38). In total, 553 participants were included in downstream analysis (Figure 2). Ethical approval was granted by No.123/EIREC2018 and No.171/EIREC2021 in Indonesia and the WEHI HREC (19/03).

**Figure 2.**
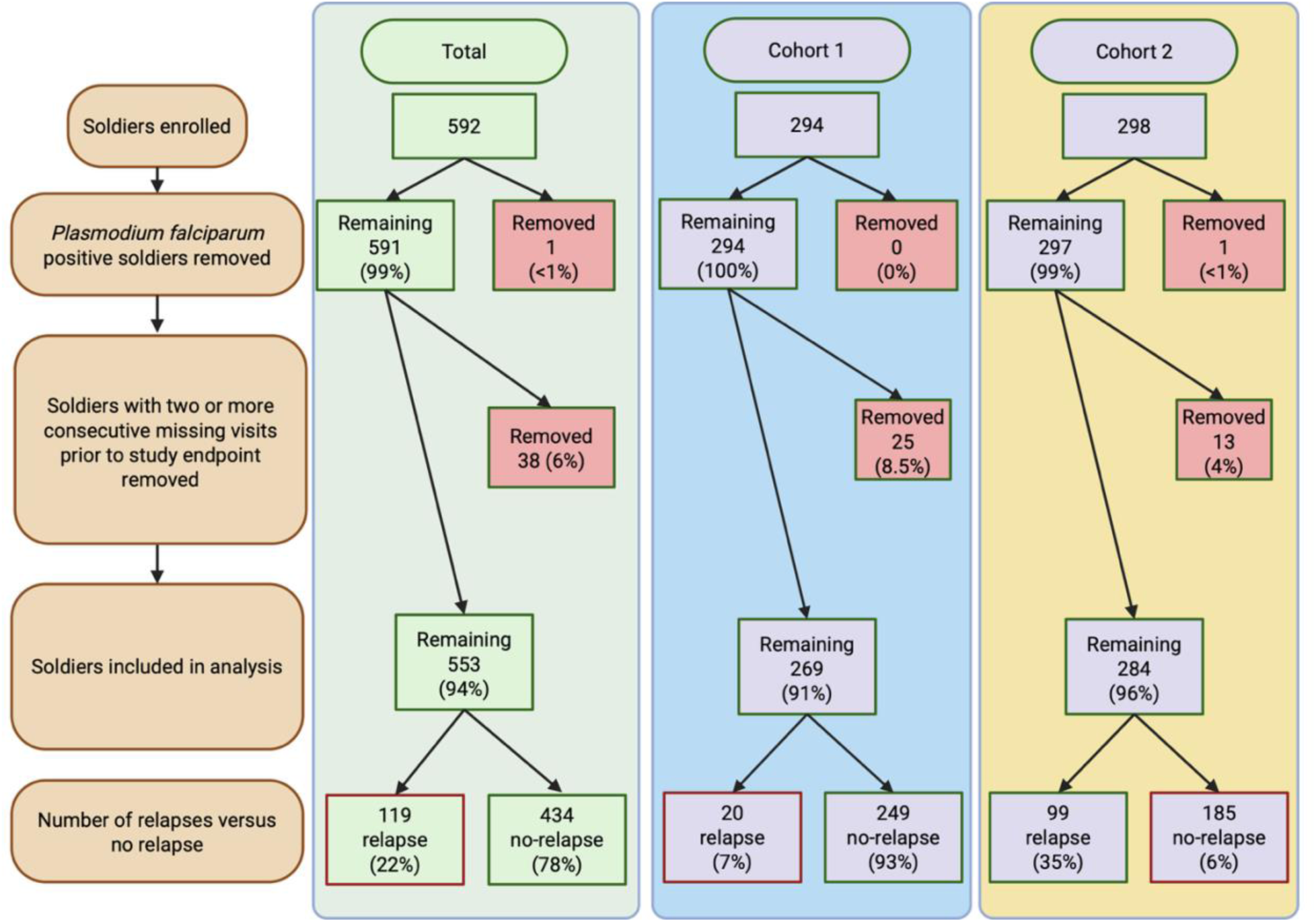
Flow chart representing soldiers included and excluded during study. Created with BioRender.com.

### Procedures

History of fever, body temperature (fever was defined as >37.5°C) and other demographic characteristics were obtained by field doctors using a structured questionnaire. Twelve ml of venous blood was collected on the day of recruitment, and then separated into whole blood, serum, packed red blood cells, and plasma in the field sites. Participants were followed up by the research team operating a 24/7 primary care clinic on the army base in cooperation with Indonesian Army medical personnel, both actively and passively, during the first six months after returning to their non-endemic/home bases. During fortnightly ACD, 500 μl of capillary blood was taken. PCD relied on the care-seeking behaviour of soldiers with malaria-compatible symptoms at the clinic on the army base, where fingerprick blood samples (500 μl) were collected for microscopy-directed diagnosis. In both cases, venous blood (5 ml) was then collected from *P. vivax-*positive soldiers. Blood was separated into pellets and plasma, and all components were stored at −80°C until usage.

### Treatment during deployment in Papua

The national guidelines recommend that soldiers who experienced a clinical *P. vivax* episode during deployment would have received unsupervised low-dose primaquine treatment of 0.25mg/kg body weight once daily for 14 days without knowing their G6PD status.

### Treatment after enrolment at the battalion

After enrolment in the study, soldiers from the first cohort were tested for glucose-6-phosphate dehydrogenase (G6PD) deficiency using the NADPH qualitative fluorescent spot test (Trinity Biologicals, USA), and soldiers from the second cohort were tested using RDT G6PD test by SD Biosensor (SD Biosensor, Republic of Korea). Both G6PD normal and deficient soldiers were eligible to participate in the study. Participants with microscopically confirmed malaria detected through PCD or ACD were treated according to Indonesian Ministry of Health guidelines. Soldiers with *P. vivax* infection received dihydroartemisinin-piperaquine phosphate (DHA-PP) for 3 days together with primaquine of 0.25 mg/kg/day for 14 days at 3.5 mg/kg total dos. Soldiers with *P. falciparum* infections were treated with three days of DHA-PP plus a single dose of primaquine (0.25 mg/kg body weight) for *P. falciparum*-infected soldiers following the Ministry of Health guideline.

### Molecular Diagnosis

DNA was extracted from packed red blood cell samples using the FavorPrepTM Blood Genomic DNA Extraction Mini Kit (Favorgen Biotech Corp, Taiwan), according to the manufacturer’s instructions. The presence of *P. falciparum, P. vivax, P. malariae,* and *P. ovale* parasites was determined by the 18S rRNA RT-PCR^15^ method using GoTaq® qPCR Master Mix (Promega Corp, Madison, WI, USA).

### Multiplexed Bead-Based Serological Assay for Antibody Measurements

Antibody levels were measured to a panel of eight selected *P. vivax* antigens coupled with magnetic beads using a Luminex MAGPIX® bead array system to all enrolment and follow-up samples as previously described.^4,16^ The eight *P. vivax* antigens used in the global algorithm were: EBP, MSP1-19, MSP5, MSP8, PTEX150, Pv-fam-a, CSS, and RBP2b (Table S1); for the dataset-specific algorithm, CSS was replaced with RAMA (Table S1). These were coupled to magnetic beads following standard methods.^16,17^

The protein-coupled magnetic microspheres was incubated with plasma from soldiers at a 1/100-dilution simultaneously with blank, positive, and negative controls. A standard curve made of two-fold serial dilution from 1/50 to 1/25,600 of positive control (hyper-immune plasma pool from Papua New Guinea) was used on each plate to allow plate-plate standardisation. Negative control plasma was from healthy individuals resided in Java, Indonesia, with no history of malaria exposures. The median fluorescent intensity (MFI) values from the MAGPIX instrument were converted to arbitrary relative antibody units (RAU) based on the standard curve data in a five-parameter logistic model packaged as an RShiny App available at: https://gitlab.pasteur.fr/tobadia/pvserotat-rshiny-app.

### Data analysis

A random forest classification algorithm was used to predict which soldiers had exposure to *P. vivax* in the prior nine months, and were thus at risk of relapse. The model used total IgG antibody responses to eight *P. vivax* antigens as predictor variables, with infection status *(recently infected and thus a risk of relapse* or *not*) as the outcome.^4,18^ The global algorithm was previously trained on antibody datasets from cohort studies in Thailand (n=680), Brazil (n=886), the Solomon Islands (n=709) and negative non-endemic controls (n=360), totalling 2635 observations. These cohorts contributed monthly qPCR-confirmed infection data. Updates to assay and algorithm since initial description^4^ are outlined in the supplementary material.

Soldiers in this study were categorised as *relapsing* or *not relapsing* based on *P.vivax* positivity by light microscopy at enrolment or during the six-month follow-up. Algorithm performance in predicting the risk of relapse was assessed using the area under the receiver operating characteristic (ROC) curve (AUC). We compared the global algorithm with a dataset-specific algorithm, trained exclusively on antibody data from the Indonesian soldier cohort (n=553). We selected a dataset-specific optimal eight-combination of antigens, and assessed the final combination using 10-fold cross validation with five repeats (methods in supplementary information).

We also examined IgG antibody differences across true positive (detected relapse, seropositive), true negative (no detected relapse, seronegative), false positive (no detected relapse, seropositive), and false negative (detected relapse, seronegative). Pairwise comparisons of log₁₀-transformed relative antibody units were performed using Welch’s t-test, assessing: (i) seronegative vs seropositive soldiers without relapse; (ii) seronegative soldiers with vs without relapse; and (iii) seronegative vs seropositive soldiers with relapse. P-values were adjusted using the Holm method.

## Results

From 553 screened participants, 119 soldiers (22%) had *P. vivax* relapse infections detected by light microscopy. Nineteen of these soldiers had a relapse detected at baseline, and were therefore treated and excluded from the six-month follow-up, as per the study protocol. Across the six-month follow-up period, 101 soldiers had a detected relapse, 69 soldiers had only one relapse, 28 had only two, and 4 had three relapses. The majority of soldiers (76/101) experience their first relapse in the first 10 weeks of the study (Figure 3).

**Figure 3.**
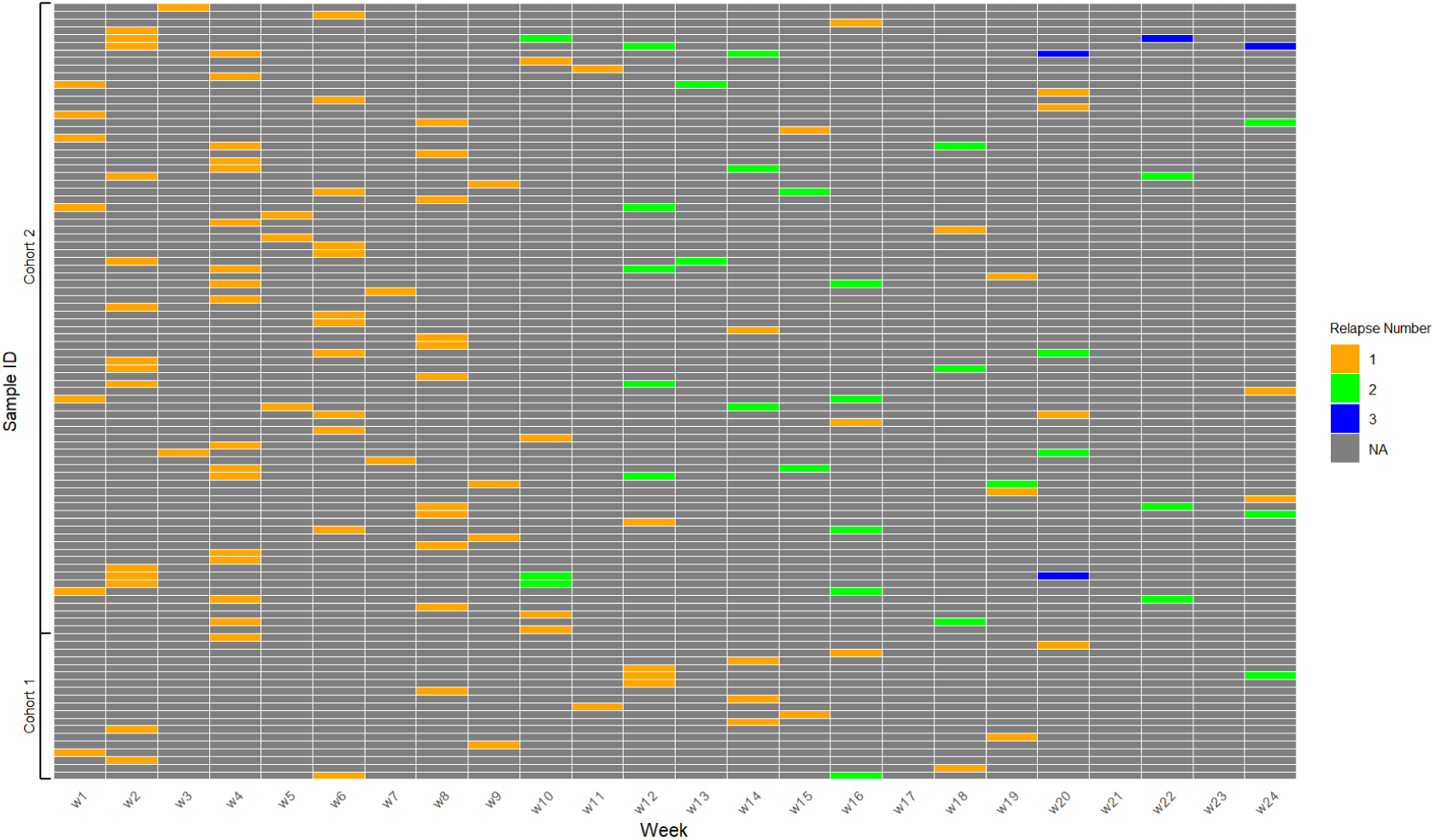
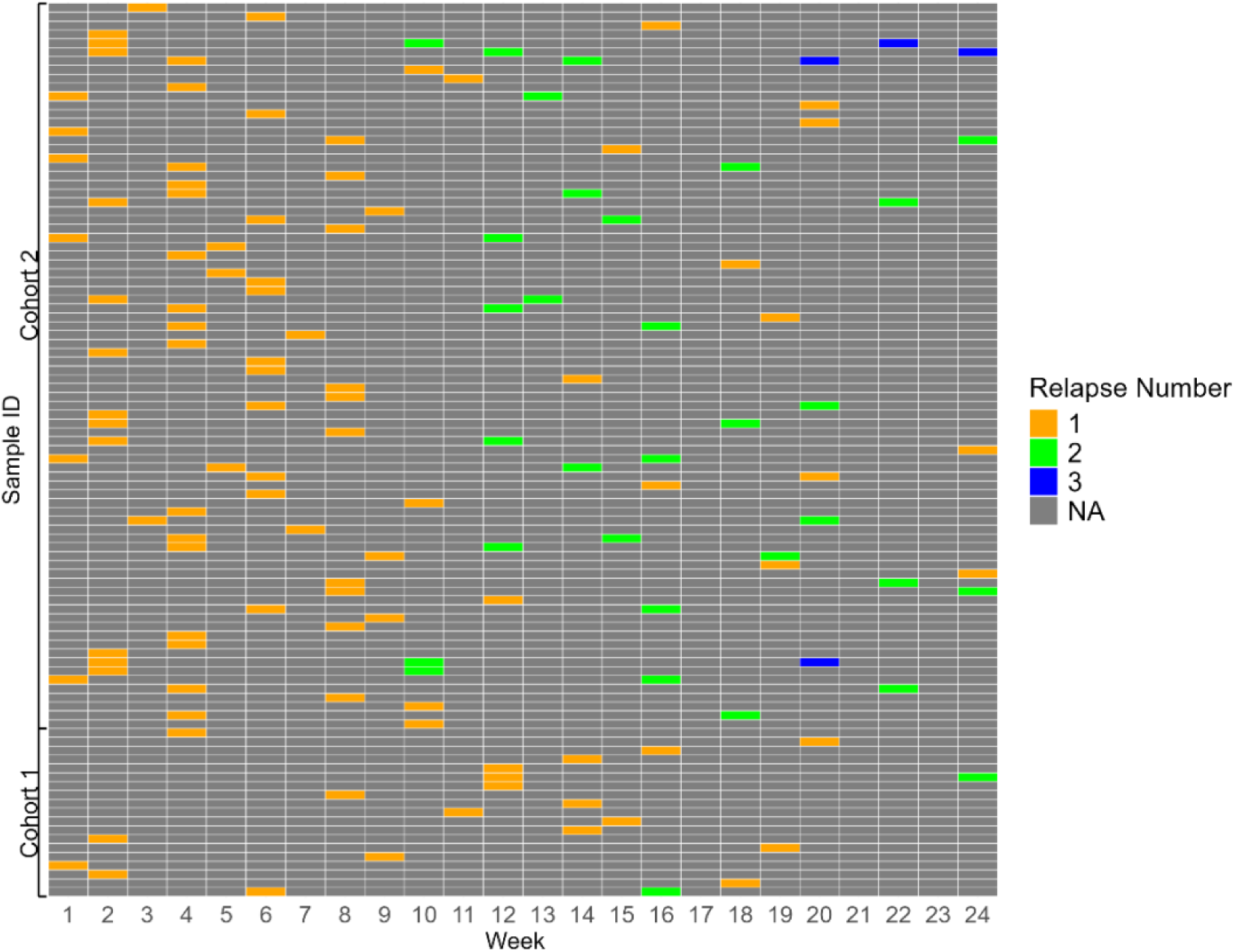
Illustration of relapse patterns during the six-month follow-up period. Active case detection by microscopy occurred fortnightly. Relapses identified through passive case detection (i.e. additional blood sampling, collected during episodes of febrile symptoms) are shown in odd numbered weeks. “NA” indicates no relapse detected by light microscopy. This figure excludes soldiers who were positive for *P. vivax* at enrolment and therefore did not participate further in the follow-up period (n=19).

There was observable heterogeneity of relapse rate between the first (20/269, 7%) and second cohort (99/284, 35%, χ^2^ = 59.908; p-value = 9.938×10^-15^). Data were analysed using both cohorts combined, but performance per cohort is provided in the supplement. The epidemiological characteristics of participants is shown in Table 1.

**Table 1.**
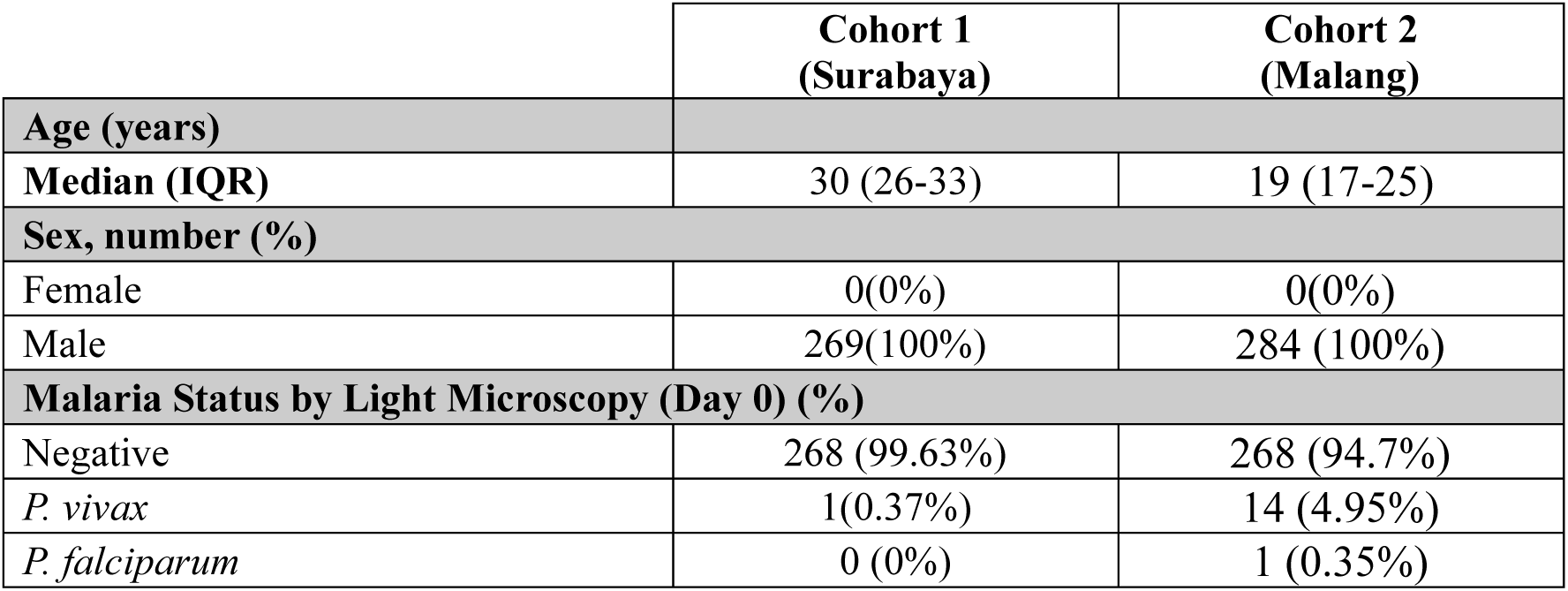
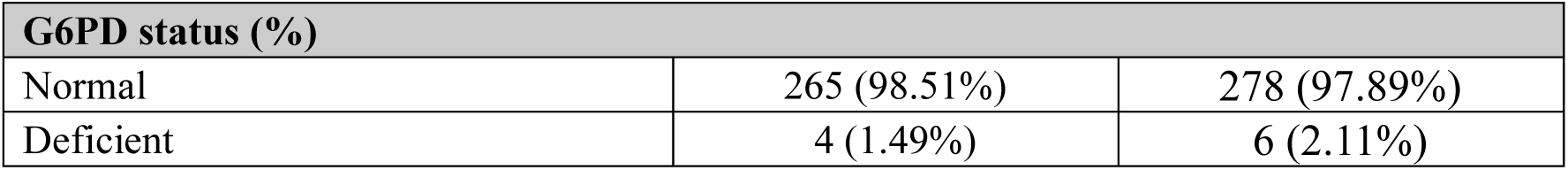
Epidemiological characteristics of participants in each soldier cohort.

|  | Cohort 1<br>(Surabaya) | Cohort 2<br>(Malang) |
| --- | --- | --- |
| **Age (years)** |  |  |
| **Median (IQR)** | 30 (26-33) | 19 (17-25) |
| **Sex, number (%)** |  |  |
| Female | 0(0%) | 0(0%) |
| Male | 269(100%) | 284 (100%) |
| **Malaria Status by Light Microscopy (Day 0) (%)** |  |  |
| Negative | 268 (99.63%) | 268 (94.7%) |
| *P. vivax* | 1(0.37%) | 14 (4.95%) |
| *P. falciparum* | 0 (0%) | 1 (0.35%) |

| G6PD status (%) |  |  |
| --- | --- | --- |
| Normal | 265 (98.51%) | 278 (97.89%) |
| Deficient | 4 (1.49%) | 6 (2.11%) |

Both algorithms performed strongly, with AUCs of 0.92 (global) and 0.93 (dataset-specific) (Figure 4A). Selecting thresholds that balanced sensitivity and specificity yielded values of 86% sensitivity and specificity for the global algorithm and 87% sensitivity and 88% specificity for the dataset specific algorithm (Figure 4A). At these thresholds, the positive predictive value (PPV) and negative predictive value (NPV) were 63% and 96% for the global algorithm, and 66% and 96% for the dataset-specific algorithm. The global algorithm correctly classified 102 out of 119 relapsing soldiers as recently infected with *P. vivax*, while the dataset-specific algorithm correctly classified 104 (Figure 4B).

**Figure 4.**
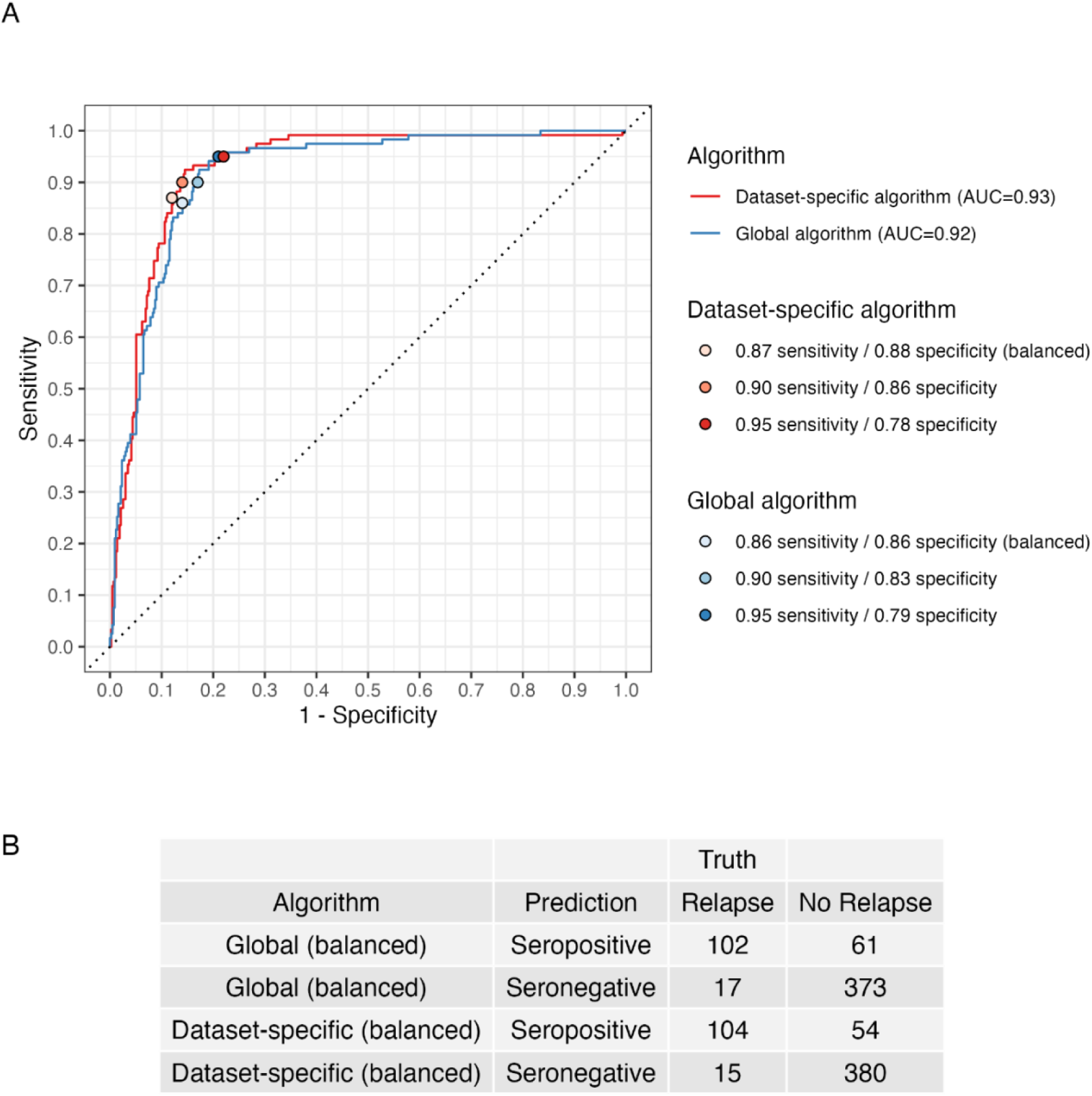
A) ROC curves for a random forest classification algorithm trained on (i) Thailand, Solomon Islands, Brazil, and malaria-naive serological data (Global algorithm) and (ii) Indonesian soldier serological data (Dataset-specific algorithm). Both algorithms were tested on the Indonesian soldier serological data. Dot-points indicate the thresholds achieving balanced sensitivities/specificities, as well as the specificities corresponding to 90% and 95% sensitivity. B) Balanced sensitivity and specificity confusion matrix for global and dataset-specific algorithms.

Total IgG antibody levels were plotted by classification status for the global algorithm (Figure 5). The true positive and true negative categories (no relapse & seronegative; relapse & seropositive) had clearly the lowest and highest antibodies to each of the 8 *P. vivax* proteins, respectively. For those with no detected relapse but seropositive, IgG antibody levels were varied but with medians statistically significantly higher than the no relapse seronegative group for all eight *P. vivax* proteins. This indicates the possibility of recent past exposure but no relapse was detected during the study follow-up period. In contrast, for those with detected *P. vivax* relapses that were incorrectly classified as not at risk, IgG antibody levels varied between the eight *P. vivax* antigens: although responses were significantly lower than in sero-positive individuals for all antigens except Pv-fam-a, antibody titres were significantly higher than in seronegative, non-relapsers for MSP1-19, MSP8, Pv-fam-a and PTEX150, but not for EBP, MSP5, RBP2b and CSS. Model estimates for pairwise comparisons are reported in S2 table and S4 figure.

**Figure 5.**
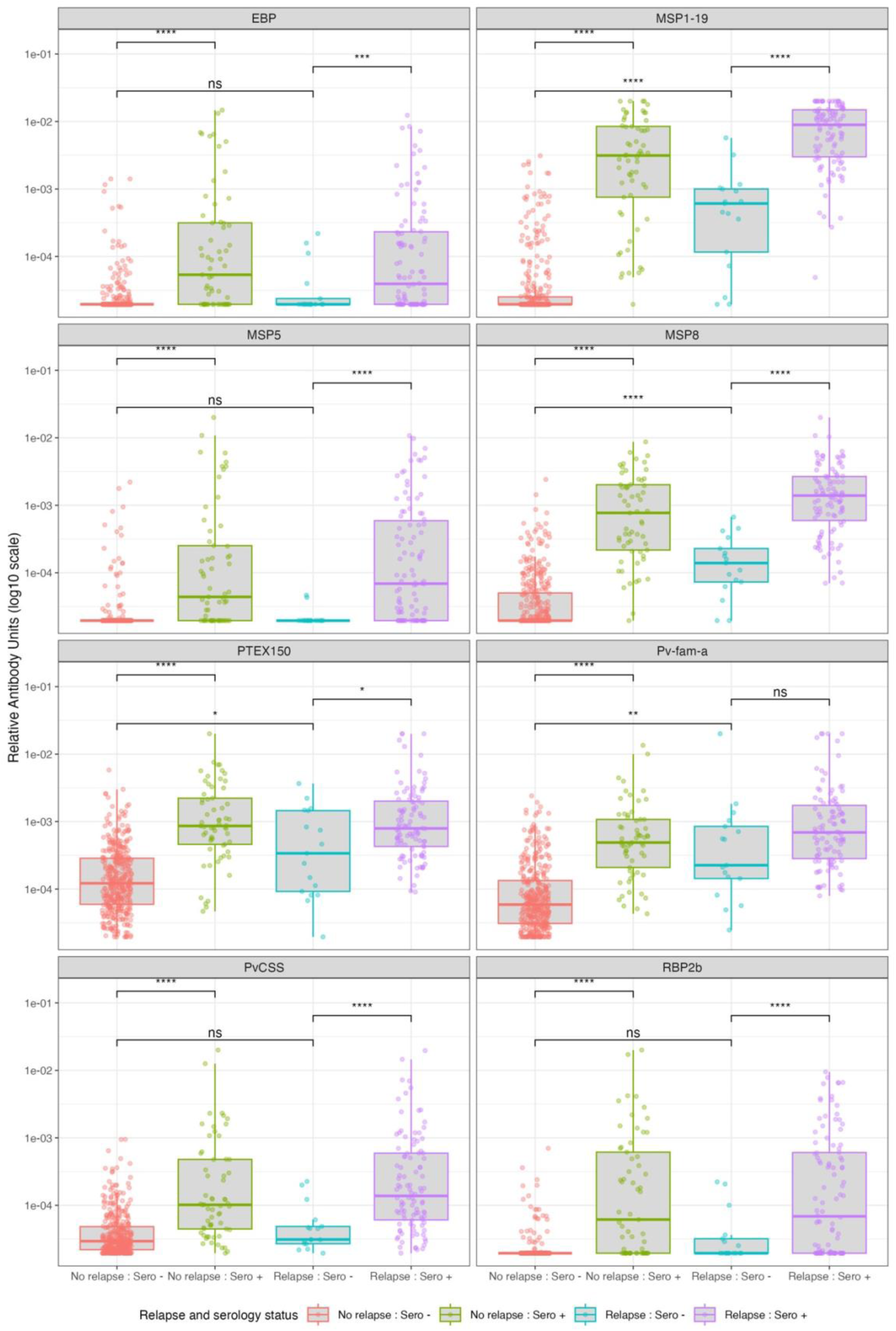
Relative antibody Unit (RAU) on log10 scale for each protein separated by classification status (seropositive or seronegative) and relapse status (relapse or no relapse) for balanced sensitivity and specificity for the global algorithm. Pairwise comparisons were performed using Welch’s t-test on log10-transformed RAU. We compared: (i) seronegative versus seropositive soldiers who had no detected relapse; (ii) seronegative soldiers with a detected relapse versus seronegative soldiers without a detected relapse; and (iii) seronegative versus seropositive individuals with a detected relapse. P-values adjusted for multiple comparisons using the Holm method.

All samples positive by light microscopy were also positive by RT-PCR. Among the microscopy-negative, sero-positive samples selected for confirmatory RT-PCR testing, eight out of 145 tested (6%) were PCR-positive. This included four out of 80 soldiers in cohort one (5%) and four out of 65 soldiers in cohort two (6%). Notably, all microscopy-negative but PCR-positive soldiers were also seropositive. No PCR-positive infections were detected among the 20% randomly selected microscopy and serology negative individuals.

The test performance results reported above use light microscopy data as the reference defining relapse status. If PCR-positive results were also included, the global algorithm AUC would increase slightly to 0.93, with a balanced sensitivity and specificity of 87%.

## Discussion

*P. vivax* remains a major hurdle for malaria elimination in the Asia-Pacific and the Americas due to the resilience against traditional control tools provided by its ability to relapse from dormant, liver hypnozoites. As indicated by the recently published WHO Preferred Product Characteristics (PPC)^19^, new diagnostic tools are urgently needed to uncover individuals with *P. vivax* liver-stage hypnozoites, who are at risk of future relapse. In this study, we explicitly demonstrate that our panel of *P. vivax* serological biomarkers, combined with a random-forest classification algorithm, identifies people at high risk of *P. vivax* relapse with high accuracy (AUC >0.92, sensitivity >86%, specificity >86%, PPV > 63%, NPV >96%). This makes our *P. vivax* serological markers of recent exposure the first ever diagnostic test that fulfils the WHO PPC for predictors of future *P. vivax* relapse PPC.

We tested the use of both a global random forest algorithm, trained on data from Thailand, Brazil and the Solomon Islands, and a dataset-specific algorithm trained and tested on the data generated from the study population in Indonesia. Their highly comparable performance demonstrates that population-specific algorithms are not required, and that a single global algorithm generalises effectively across diverse geographic regions. This builds on previous work where the *P. vivax* serological exposure markers have been tested for their ability to predict recent past exposure and/or generate epidemiologically expected results in multiple countries including Peru^20,21^, Cambodia^22^, Indonesia^23^, and Thailand^18^. Furthermore, the *P. vivax* antigens underlying these serological markers have recently been assessed for levels of genetic diversity^24^, with most having minimal polymorphisms and limited evidence of geographic structure. Although some Indonesian soldiers who relapsed during the study were classified as seronegative, this is unlikely to be related to antigen genetic diversity. These individuals displayed low antibody responses across the antigens EBP, MSP5, RBP2b, CSS and RAMA, all of which have low levels of global genetic diversity except MSP5. Diversity in MSP5 is not geographically structured. The misclassification may instead be due to very recent *P. vivax* infections with insufficient time for antibody response to develop, or other individual-level variables that may impact strength of antibody responses. Overall, sensitivity was still exceptionally high at 86%, exceeding that of the original validation study, where 80% was reported.^4^ Whilst this performance may appear limited in comparison to other diagnostic tests, it must be considered in context. The random forest is tasked with distinguishing recent *P. vivax* infection (within the prior nine months, when most individuals would have relapsed), from older *P. vivax* infections. This challenging classification highlights the robustness of the algorithm.

Some misclassification is expected. False positives may arise from either incorrect underlying truth (i.e. soldiers had no detected relapses by microscopy but may have had undetected *P. vivax* infections) or incorrect prediction based on the serology. Additionally, soldiers experiencing a clinical *P. vivax* episode during their deployment in Papua, may have been administered primaquine in accordance with national treatment guidelines. However, no data was available on which soldier had received radical cure prior to enrolment and whether they may have completed the full course of treatment. In such cases, they would have been correctly identified as at risk of relapse (seropositive), but would have had no detected relapse during follow up. Unsupervised, low-dose primaquine treatment has been reported to reduce relapse risk by only 10%^25^, which could partially explain some of the 59 soldiers in the no relapse, seropositive (false positive) group. Conversely, a small number of relapsing individuals may have been missed by light microscopy; however only 6% of light microscopy-negative soldiers were RT-PCR positive and all of these were seropositive. PCR did not detect any additional infection among 20% of seronegatives indicating that few relapses were missed in this study. As relapse risk in this study was defined solely by light microscopy, the reported performance metrics reflect classification against this reference standard. We performed an exploratory analysis where we included any PCR-positive, light microscopy-negative samples as relapses, which marginally increased the AUC to 0.93. This indicated improved apparent performance of the sero-diagnostic test. Taken together, this suggests that light microscopy–based relapse detection provides a conservative (lower-bound) estimate of test algorithm performance, while analyses incorporating PCR reflect the upper bound.

This study is uniquely positioned to evaluate relapse prediction, due to its access to a cohort of individuals naturally exposed in Papua and then relocated to a malaria-free area, allowing clear attribution of subsequent infections to relapses rather than newly acquired infections. Although relapse proportions differed between the two battalions (reflecting variation in exposure risk; (Figure 1), the global algorithm performed consistently in each cohort (AUC>0.88, supplementary information), demonstrating robust and consistent classification performance. However, there are limitations to this study design, namely that only men were included simply due to the demographics of this soldier population in Indonesia. The global algorithm was trained on datasets including both men and women, limiting the potential impact, however it means the algorithm has not been validated on women for identifying risk of relapse (but has been for risk of recent blood-stage *P. vivax* infection). Likewise, the soldiers in this study were all adults aged between 18-65 years, however the global algorithm was trained on datasets that included children. Another limitation is that the study period was six-months. Some soldiers may have gone on to relapse after this period, meaning they would have been incorrectly classified as false-positive, underestimating performance. Finally, a more balanced dataset in terms of the proportions of soldiers who relapsed would have provided a better assessment of the algorithms classification performance. Overall, these limitations largely signify that the observed sensitivity and specificity of our serological test for detecting risk of relapse may be underestimated.

At a 86% sensitivity and specificity for detection of future relapse risk, this indirect biomarker-based serological test will be a powerful addition to the arsenal required for global elimination of *P. vivax* malaria. At this performance it forms the foundation of *P. vivax* serological testing and treatment (*Pv*SeroTAT), a targeted intervention designed to reduce relapse-driven transmission.^4^ The laboratory-based Luminex assay used here aligns with the PPC characteristics put forth for use of serology for surveillance, and is already being applied in pilot *Pv*SeroTAT^26^ and sero-surveillance studies.^27^ Modelling suggests that *Pv*SeroTAT could reduce *P. vivax* transmission by 50-80%^28,29^, particularly in low-transmission settings, while using up to 80% less drug treatments than the alternative, mass drug administration, thus reducing costs and improving safety. This makes serological identification (and treatment) of individual at risk of relapse, the first new public health intervention aimed directly at eliminating the hidden *P. vivax* hypnozoite reservoir.

## Data Availability

All data produced in the present study are available upon reasonable request to the authors

## Acknowledgments

We would like to extend our gratitude to the Chief of the Indonesian Army Republic of Indonesia and his staffs, Indonesian Army Medical Centre, and Eijkman Institute for Molecular Biology. We acknowledged proteins produced and purchased from the Tham Laboratory (Wai-Hong Tham, WEHI, Melbourne, Australia), the Cowman Laboratory (Pailene Lim, Stephen Scally, WEHI, Melbourne, Australia), the WEHI Protein Production Facility (Marija Dramicanin, Melbourne, Australia), CellFree Sciences (Yokohama, Japan), and ZiP Diagnostics (Melbourne, Australia). We acknowledge efforts of both Natalie Senzo and Julie Healer as project managers at WEHI. We would also like to acknowledge the contribution of Dedi Sudiana from Oxford University Clinical Research Unit, Jakarta, Indonesia for site setup. The authors acknowledge the Victorian State Government Operational Infrastructure Support and Australian Government NHMRC IRIISS.

## Author contributions

Conceptualisation: RN, MTW, RJL, JKB, IJM. Field-work and co-ordination: LT, LLE, NF, FAN, HH, PPK, ANF, GH, MC, TAK, DS, SS, FW, NN, DS, WB, YE, MDW, FF. Laboratory work: RASU, RA, ARA, AMP. Data curation: RASU, LS, ES, ARA, NN, RJL. Formal analysis: LS. Investigation: RN, RASU, LS, ES, ARA, NN, MTW, RJL, JKB, IJM. Resources: RM, RJL. Supervision: RN, RJL, JKB, IJM. Visualisation: LS. Writing – original draft: RN, LS, ES, ARA, MAH, LJR, RJL, IJM. Writing– review & editing: all authors.

## Supplementary information

### PvSeroTAT algorithm

Since publication of our sero-diagnostic assay^4^, we have made several improvements to both our assay protocol^16^ and the machine learning classification algorithm, namely (i) the optimisation of our selection of antigens that are associated with recent exposure, (ii) generation of a new training dataset of antibody responses towards our panel of antigens using magnetic beads (same samples as previously), and (iii) further optimisation of our random forest classification algorithm through hyperparameter tuning (Smith et al in preparation). Below we outline these details.

### Details of training of global algorithm

We consider 10 *P. vivax* antigens (Table S1) in our panel (details of downselection process outlined ^4^). We created a random forest classification algorithm for all possible combinations of eight antigens from the 10-antigen panel, using 10-fold cross validation with five repeats. We use the top-performing combination of eight antigens, evaluated using the receiver operating characteristic (ROC) curve (AUC).

**Table S1:** P. vivax proteins utilised in the global and dataset-specific algorithms. GeneID from either PlasmoDB or GenBank.

| Antigen name | Gene ID | Sequence region (amino acid length) | Antigen source | Algorithm |
| --- | --- | --- | --- | --- |
| EBP | KMZ83376.1b | 109-432 (324) | WEHI Protein Production Facility | Global; dataset-specific |
| PTEX150 | PVX_084720 | 24-908 (885) | CellFree Sciences | Global; dataset-specific |
| Pv-fam-a | PVX_096995 | 56-480 (425) | ZiP Diagnostics | Global; dataset-specific |
| MSP5 | PVX_003770 | 25-364 (340) | ZiP Diagnostics | Global; dataset-specific |
| RBP2b | PVX_094255 | 161-1454 (1294) | WEHI Tham Lab | Global; dataset-specific |
| MSP1-19 | PVX_099980 | 1623-1715 (93) | ZiP Diagnostics | Global; dataset-specific |
| CSS | PVX_086200 | 22-381 (360) | WEHI Protein Production Facility | Global only |
| MSP8 | PVX_097625 | 24-463 (440) | WEHI Protein Production Facility | Global; dataset-specific |
| RAMA | PVX_087885 | 634-709 (76) | ZiP Diagnostics | Dataset-specific only |
| S16 | PVX_000930 | 31-110 (80) | CellFree Sciences | Not selected in either. |

We train the final random forest algorithm using 10,000 trees. We optimised the hyperparameters (i) number of predictors randomly sampled at each split and (ii) tree stopping criteria minimum number of nodes using Bayesian optimisation. This resulted in two predictors randomly sampled at each split and tree stopping criteria set to seven in the **ranger**^30^ package in R version 4.4.1.^31^ This classification model was trained on all data from the longitudinal cohort studies, plus negative controls (malaria endemic samples: Thailand n=680, Brazil n=886, the Solomon Islands n=709; negative non-endemic controls: Australian Red Cross = 97, Rio Negatives = 96, Thai Red Cross = 69, Victorian Blood Donor Registry = 98, totalling 2635 observations). This random forest, trained on the longitudinal cohorts and negative controls, was then used to classify the Indonesian soldier cohorts as *recently infected* or *not recently infected*. To present balanced sensitivity and specificity, we determined the random forest votes threshold where sensitivity and specificity are balanced (i.e. where lines cross in figure S1). Similarly, we determined which random forest votes threshold related to a sensitivity was 90 and 95%, and presented the corresponding specificity.

### Details of training and testing of dataset-specific algorithm

To compare the classification performance of the global algorithm to a dataset-best (i.e. the dataset-specific algorithm; a random forest trained and tested specifically on the Indonesian soldier antibody levels), we determined the optimal 8-antigen combination by creating all possible combinations of eight from the 10-antigen panel. We created a random forest algorithm for each possible combination, using 10-fold cross validation with 5 repeats. We evaluated classification performance using AUC.

As with the global algorithm, we used 10,000 trees and optimised the hyperparameters (i) number of predictors randomly sampled at each split and (ii) tree stopping criteria using Bayesian optimisation for the dataset-specific algorithm. The optimal parameters were seven and two respectively. This final random forest was trained and tested on soldier dataset. We present the mean results for 10-fold cross validation.

**Figure S1.**
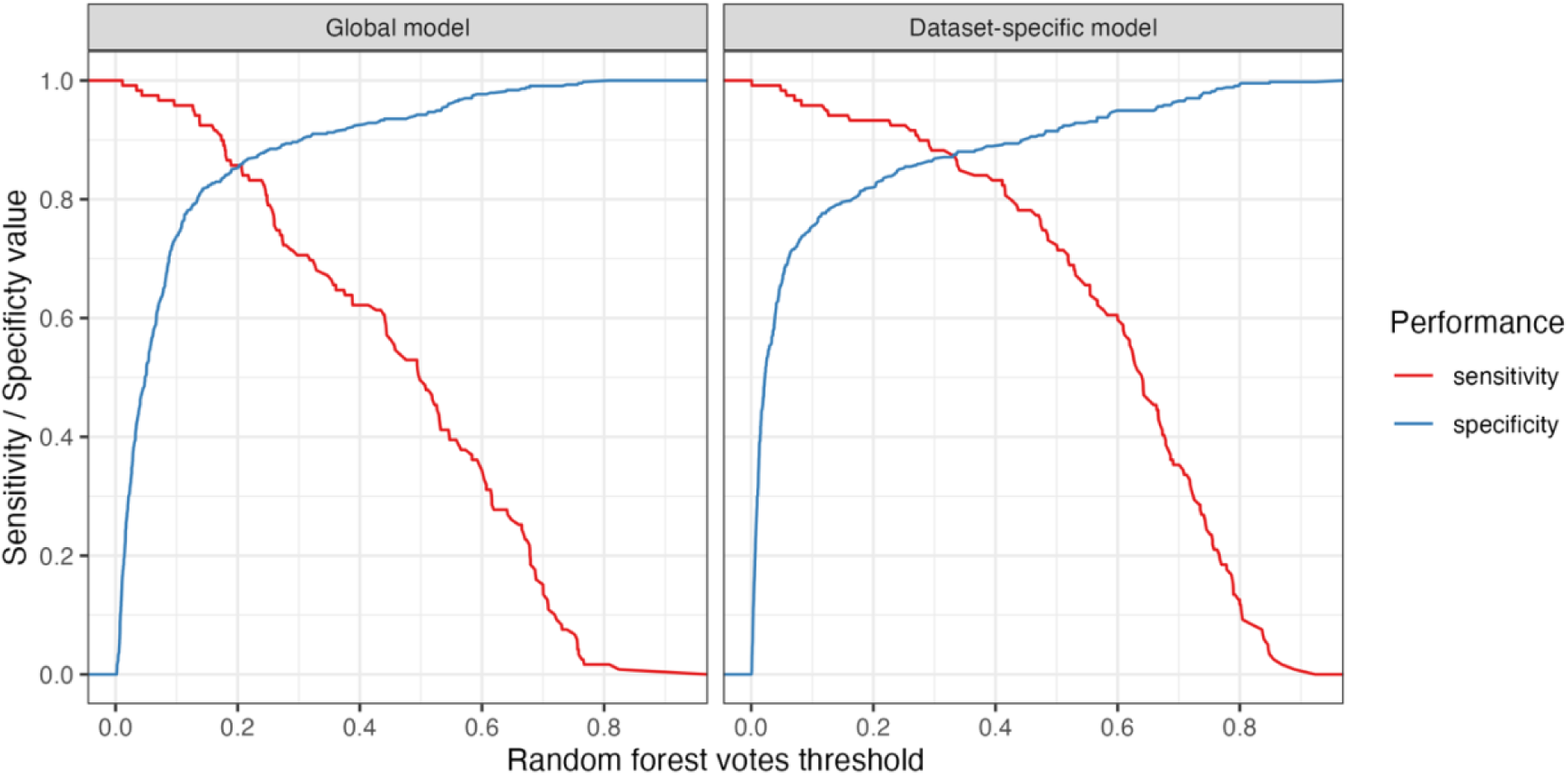
Relationship between sensitivity and specificity for both random forest algorithms. When sensitivity and specificity are balanced (i.e. optimised for the highest possible values of sensitivity and specificity, where the lines intersect), the global algorithm had a sensitivity of 0.86 and specificity of 0.86 and a positive predictive value of 0.63 and a negative predictive value of 0.96. The dataset-specific algorithm had a sensitivity of 0.87 and specificity of 0.88 and a positive predictive value of 0.66 and a negative predictive value of 0.96.

### Global algorithm performance on separate cohorts

The global algorithm had an AUC of 0.93 for soldier cohort 1 and 0.88 for soldier cohort 2 (see Figure S2(A) for corresponding ROC curves).

**Figure S2:**
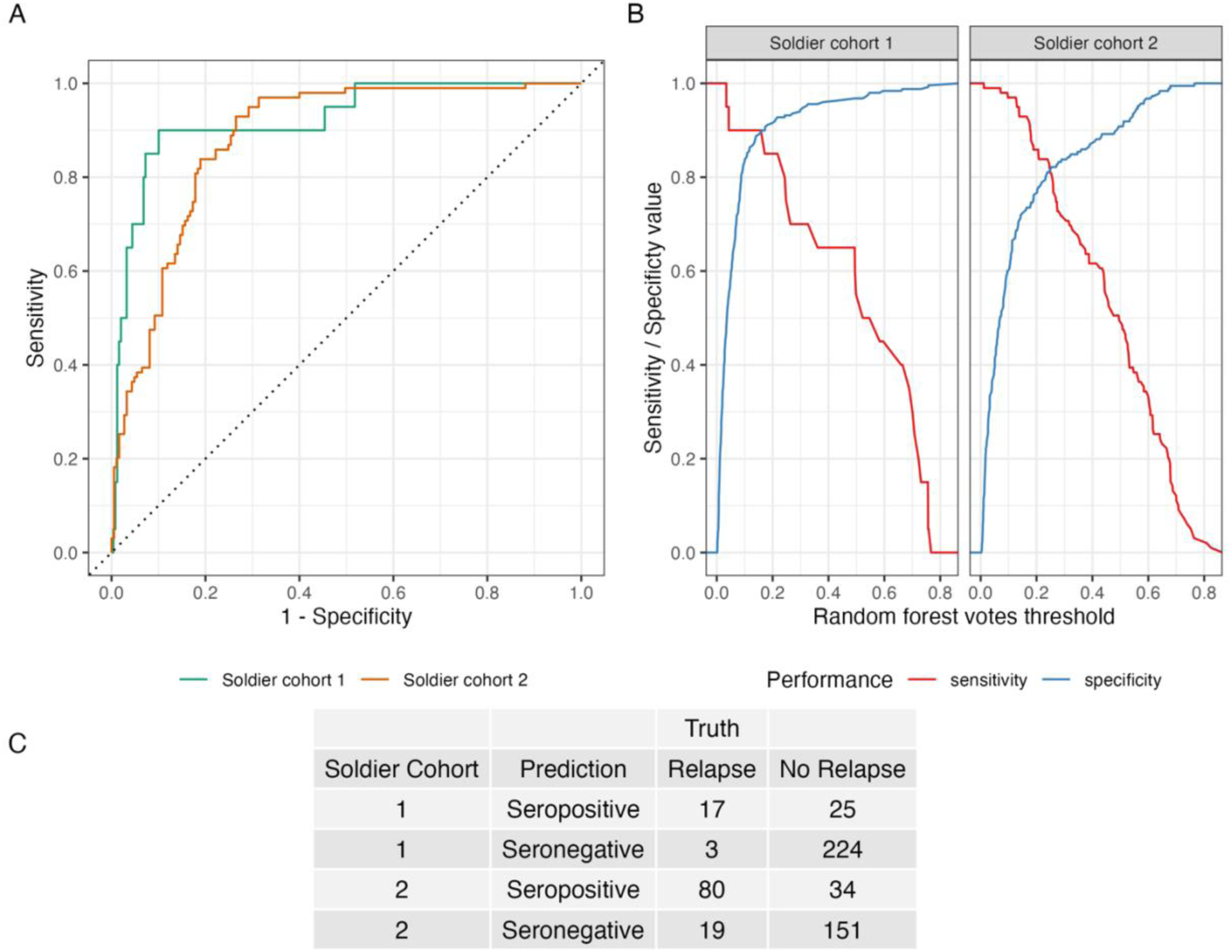
A) ROC curves for a random forest classification algorithm trained on Thailand, Solomon Islands, Brazil, Australia serological data and tested on Indonesian soldier cohort 1 and soldier cohort 2 serological data. B) Relationship between sensitivity and specificity for soldier cohort 1 and 2, showing the relationship when varying random forest model votes threshold between 0 and 1. C) Balanced sensitivity and specificity confusion matrix for soldier cohort 1 and 2.

Figure S2B shows the relationship between sensitivity and specificity for both soldier cohorts When sensitivity and specificity are balanced (i.e. optimised for the highest possible values of sensitivity and specificity, where the lines intersect in Figure S2B), the global algorithm applied to soldier cohort 1 had a sensitivity of 0.85 and specificity of 0.90, and a positive predictive value of 0.41 and a negative predictive value of 0.99. Soldier cohort 2 had a sensitivity of 0.81 and specificity of 0.82, and a positive predictive value of 0.70 and a negative predictive value of 0.89. Figure S2C presents the confusion matrix per cohort.

### Linear model estimates and predicted effect sizes

**Figure S3.**
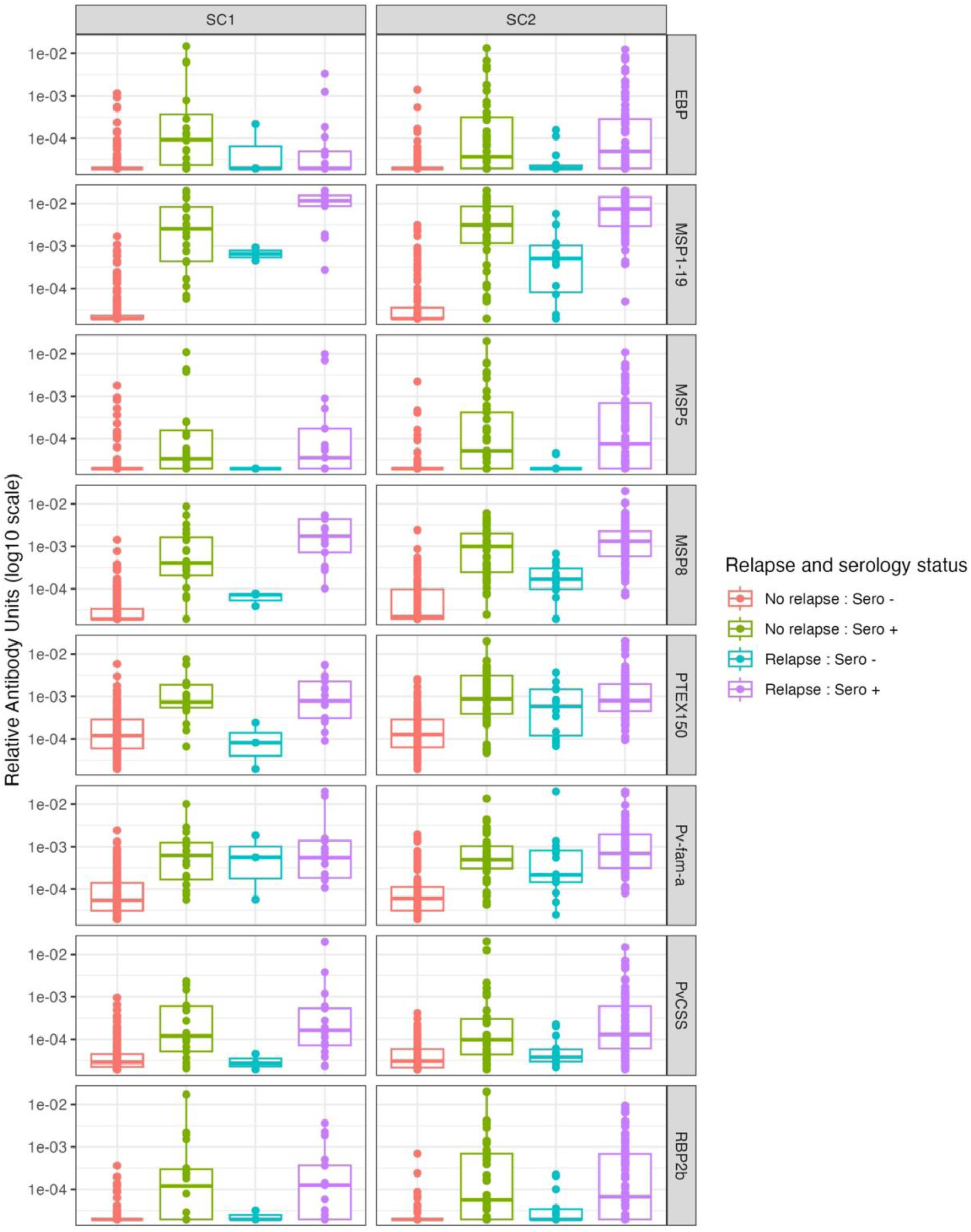
Boxplots illustrating the median and 25^th^ and 75% percentiles of the relative antibody units on log10 scale for each antigen, separated by cohort (SC1 = soldier cohort 1, SC2 = soldier cohort 2) and relapse and serology status.

**Table S2.**
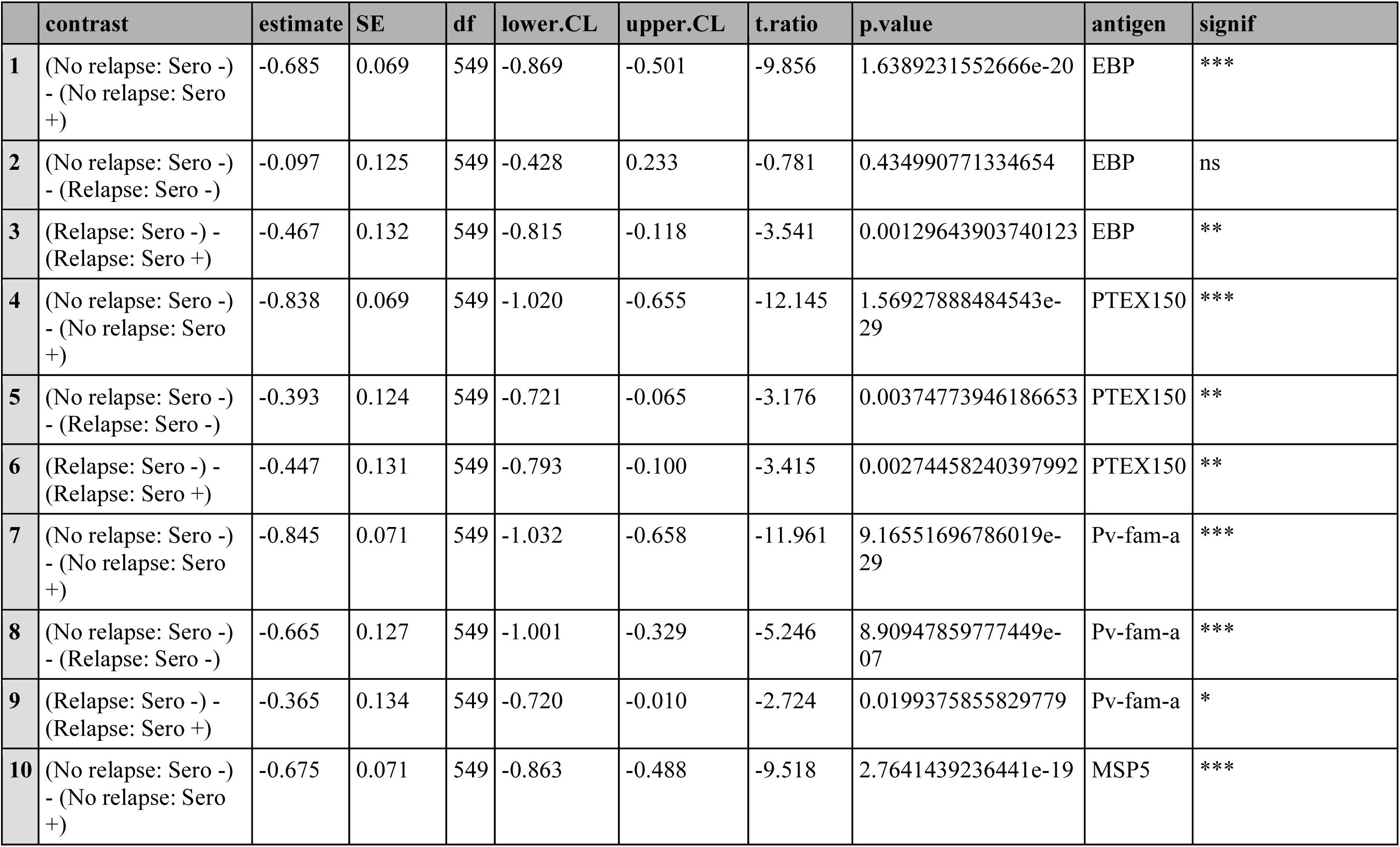
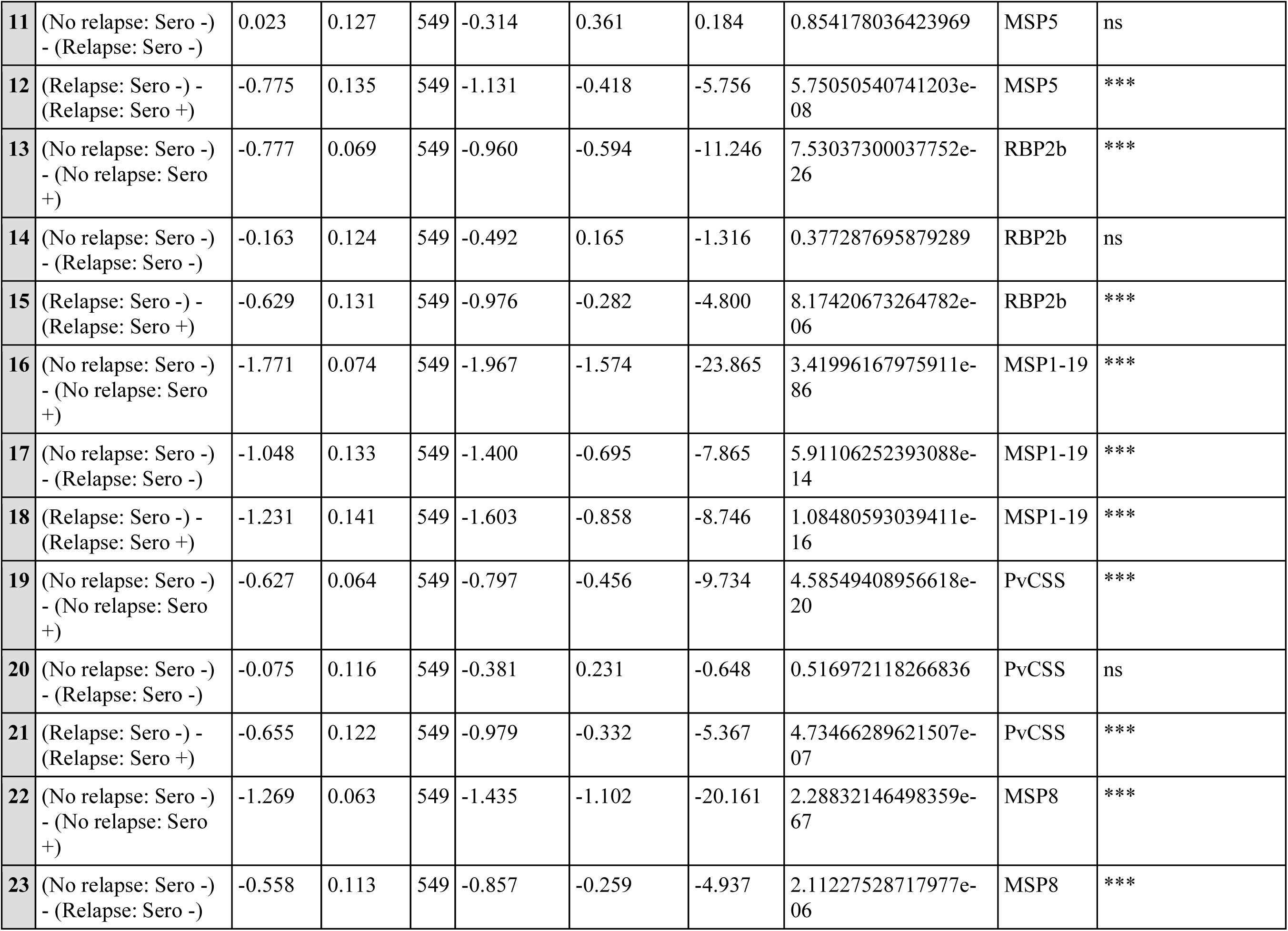
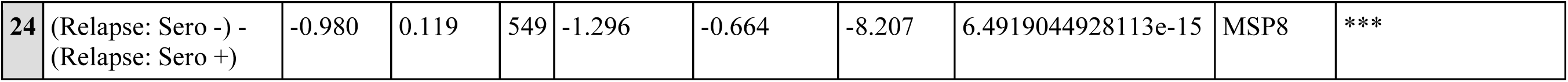
Estimated contrasts from linear models of log10-transformed RAU by relapse-serostatus category. Models were fitted separately for each antigen with relapse-serostatus category as the predictor. Reported contrasts correspond to the pre-specified pairwise comparisons, with 95% confidence intervals and Holm-adjust p-values.

**Figure S4.**
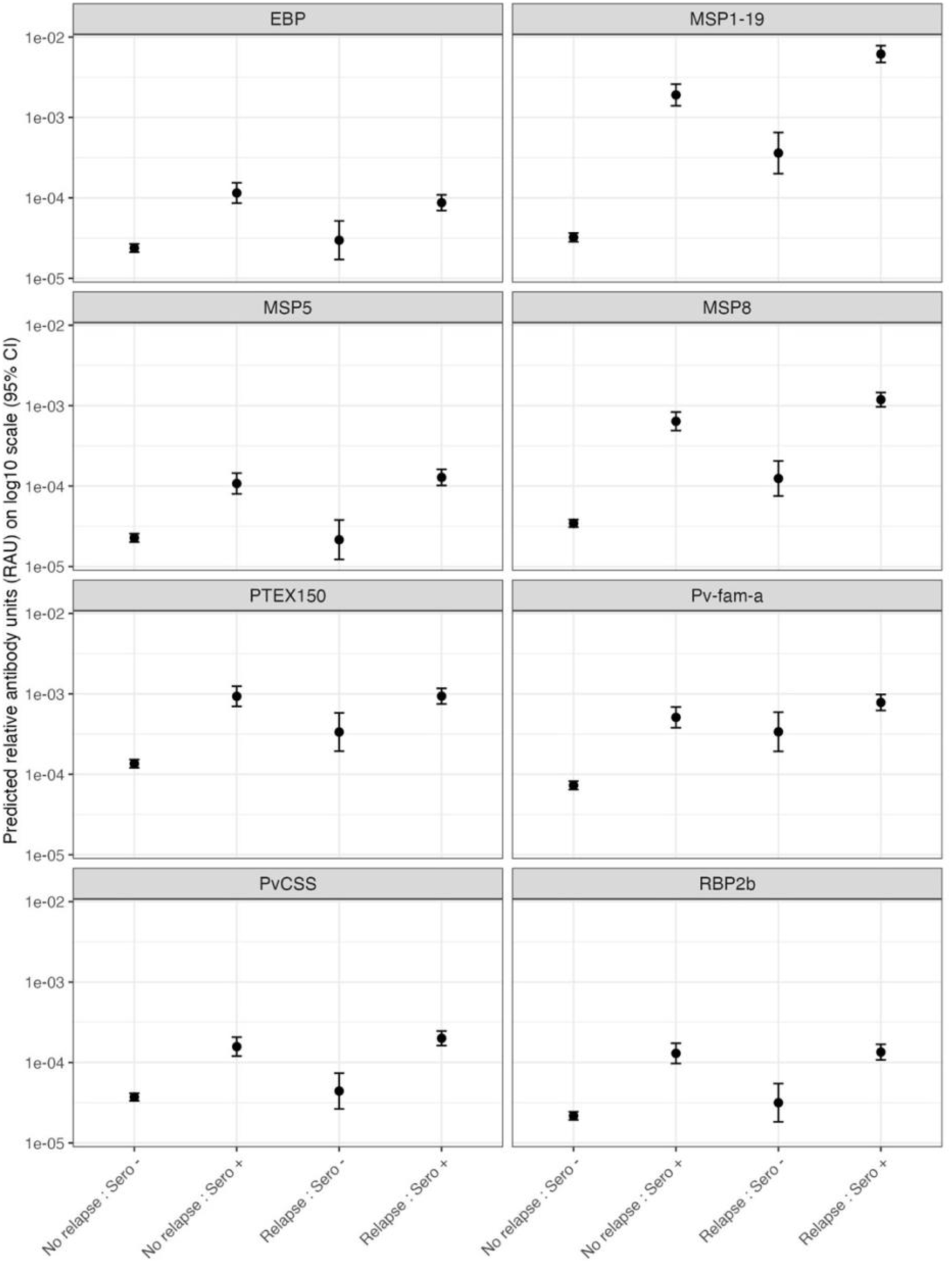
Model predicted estimates of IgG antibody responses by relapse-serostatus category. Points represent estimated log10 estimates in relative antibody units from linear models fitted separately for each antigen, with bars indicating 95% confidence intervals.

